# Dynamic landscape of mpox importation risks driven by heavy-tailed sexual contact networks among men who have sex with men in 2022: a mathematical modeling study

**DOI:** 10.1101/2023.10.06.23296610

**Authors:** Sung-mok Jung, Fuminari Miura, Hiroaki Murayama, Sebastian Funk, Jacco Wallinga, Justin Lessler, Akira Endo

## Abstract

**Background:** The 2022 global mpox outbreak spread rapidly, primarily among men who have sex with men in Western countries. The number of countries with new mpox importation events quickly rose in the early phase of the outbreak, but the rate of increase subsequently slowed down, having many countries without any reported cases.

**Methods:** We developed a mathematical model of international dissemination of mpox infections that accounts for heterogeneous sexual contact networks, infection-derived immunity in the network, and global mobility of infectious individuals. We used this model to characterize the mpox importation patterns observed in 2022 and to discuss the potential of further international spread.

**Findings:** Our analysis showed that the proposed model better explained the observed importation patterns than models not assuming heterogeneity in sexual contacts. Estimated importation hazards decreased from August 2022 in most countries, faster than the decline in the global case counts. We assessed each country’s potential to export mpox cases until the end of an epidemic in the absence of interventions and behavior changes, identifying countries capable of contributing to future international spread if they experience sustained local transmission.

**Interpretation:** Our study suggested that the accumulation of immunity among high-risk individuals over highly heterogeneous sexual networks may have contributed to the slowdown in the rate of mpox importations. However, our analysis identified the existence of countries still with the potential to contribute to the global spread of mpox, particularly those with sizable susceptible populations and large international travel volume. This highlights the importance of ensuring equitable access to treatments and resources to prevent the global resurgence of mpox.

**Research in context:** *Evidence before this study:* Mathematical models incorporating interconnectivity between countries have been used to assess the risk of international disease introductions. These approaches have assumed homogenously mixing local populations and have been successful in capturing the international importation patterns observed in previous global respiratory infection outbreaks such as influenza and COVID-19. However, it remains uncertain whether these models can be extended to the context of mpox, which has been transmitted predominantly through sexual activities among men who have sex with men. Previous studies have underpinned the significant role of infection-derived immunity in highly heterogeneous sexual networks in slowing down its transmission. Such key characteristics of mpox have not yet been incorporated in existing approaches to assessing the risk of international spread, which may lead to misguided public health decision-making.

*Added value of this study:* We constructed a mathematical model of international dissemination of mpox infections considering heterogeneous sexual networks and infection-derived immunity, as well as the global mobility of infectious individuals. By applying this model to the observed mpox importation patterns in 2022, we found that incorporating the accumulation of immunity among high-risk individuals better explains the observed slowdown in the rate of mpox importations between countries. Additionally, the model highlighted the presence of countries that still have the potential to contribute to the global spread of mpox, particularly those with large susceptible populations and a high volume of international travel.

*Implications of all the available evidence:* Our study adds to the growing evidence on the role of accumulated immunity among high-risk individuals in the slowdown the mpox transmission in the 2022 outbreak, which we found to be also the key to better understanding the global importation patterns. Without incorporating this effect, existing modeling approaches may overestimate the mpox importation risks, especially from countries where high-risk groups have already acquired immunity early in the outbreak. Furthermore, our visualization of large susceptible populations remaining in less affected countries, including low- and middle-income countries, highlights the importance of assessing the onward risk of case importation and ensuring equitable access to treatment and control measures in these at-risk countries.

## Introduction

The global mpox outbreak emerged in May 2022 and spread predominantly among men who have sex with men (MSM). The dominant mode of transmission in this outbreak is considered to be direct contact associated with sexual activities^1^, which is in stark contrast with previous outbreaks driven by animal-to-human or household transmissions^2,3^. Although there was a rapid surge of cases in the initial phase of the outbreak in affected countries in Europe and the Americas, they saw a declining trend in incidence starting in early August 2022^4^. While this decline may be partly attributable to interventions such as vaccination campaigns and voluntary behavior changes among high-risk populations^5^, several studies suggested that infection-derived immunity, especially among individuals who have many sexual partners, could explain the observed peaks^6,7^.

The 2022 mpox outbreak shows a unique international spreading pattern, distinct from previous global respiratory infection outbreaks (e.g., SARS, MERS, H1N1 pandemic influenza, and COVID-19)^8–10^. The first case was reported in the UK on 7 May, followed by identifications in a number of previously non-endemic countries in Europe and North America in mid-May, and later in other regions. However, despite its rapid spread in the initial phase, there was a noticeable slowdown in the number of countries that have experienced their first imported case following the decline in global cases in August 2022, and a considerable number of countries in Asia and the Middle East had not seen importations by the end of 2022^11^. Such rapid saturation of first importation events was not observed in the COVID-19 pandemic, where almost every country imported cases in the first few months (**Fig-S1**), although these two infections were estimated to have comparable initial reproduction numbers^12,13^. As a consequence, there remain countries where a large fraction of the population is susceptible to mpox—this may have contributed to the current upsurge in mpox cases in China, raising concerns about the potential resurgence of global mpox cases centered in Asia^14,15^.

Understanding the mechanism and risk of disease introductions between populations provides countries with timely situational awareness during a global outbreak. To this end, mathematical modeling approaches incorporating connectivity between countries have been widely used. The process of case importation is often modeled using proxies of travel volume including international flight data^16–18^ or more granular data accounting for geographical proximities^10,19^. While such approaches often simplified local-scale transmission dynamics by explicitly or implicitly assuming homogeneous mixing between individuals, they succeeded in capturing the observed importation pattern in the previous global respiratory infection outbreaks^8,9,20^. However, it has not been established whether such models can also be applied to infections primarily transmitted through sexual activities, in which greater individual-level variation than other forms of contact (typically represented by heavy-tailed distributions of sexual partners) is known to exist^21,22^. In such highly heterogeneous sexual contact networks, infection selectively spreads among individuals with many sexual partners, and the susceptible population in this higher-risk group is rapidly depleted by infection-derived immunity (“selective depletion of susceptibles”^23,24^). This selective transmission process could cause a drastic shift in the transmission potential of infectious individuals over time; that is, infectious individuals at an earlier phase of the epidemic typically have more chances of transmission (and hence contribute to international spread if the corresponding parts of the sexual network share the same characteristics) than those in a later phase. This shift may explain the unique importation patterns of the current mpox outbreak (rapid but limited geographical spread) as opposed to those observed in the COVID-19 pandemic.

Conventional case importation models assuming homogeneous mixing may not be an ideal tool for an outbreak where sexual activities play a dominant role. To better understand the global importation patterns of mpox, we developed a mathematical model of international case importations accounting for selective depletion in highly heterogeneous sexual networks. We applied this model to describe the importation patterns in 2022 and retrospectively estimate the potential of mpox case exportation.

## Materials and Methods

### Data source

The incidence data of mpox cases by date of reporting and symptom onset was retrieved from the World Health Organization (WHO) website^4^, from 7 May (the reporting date of the first case in the UK) through 1 October 2022 (up to which daily incidence was available in most countries). When symptom onset dates were unavailable, they were imputed using the reporting delay distribution (see *Supplementary Materials*). Countries with mpox cases prior to the current global outbreak (i.e., Cameroon, Central African Republic, Democratic Republic of the Congo, Ghana, Liberia, Nigeria, the Republic of the Congo; Ghana reported no human cases but the virus was found in animals)^25^ were excluded from the analyses given the uncertainty in the role of MSM populations in the mpox dynamics in these countries and their relatively small case counts. The estimated MSM population sizes were collected from the Joint United Nations Programme on HIV/AIDS (UNAIDS) dashboard and report^26,27^, which were assumed to represent the at-risk population in each country. If unavailable, the estimate was imputed using the subregional median of the MSM proportion (following the 17 subregions in the UN geoscheme^28^). International travel volume was obtained from the World Tourism Organization (UNWTO) 2019 outbound tourism data^29^ (more details are described in *Supplementary Materials*).

### Risk of mpox importation

The time-varying hazard of importing the first mpox case in each country was modeled assuming that importation events represent residents who acquired an infection while traveling to a ‘source’ country. The expected number of new secondary mpox cases generated by local cases in source country 𝑗 at time 𝑡 (i.e., discrete time with the unit of day), 𝐺_*j*_(𝑡), is expressed as

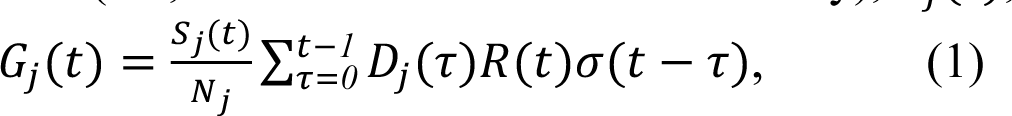

where 𝑆_*j*_ and 𝑁_*j*_ are respectively the susceptible and total population sizes, 𝐷_*j*_(𝑡) is the daily incidence by symptom onset date, and 𝜎(⋅) is the probability mass function of the serial interval^30^. 𝑅(𝑡) is the reproduction number, which is defined as the average number of secondary transmissions from cases infectious at time 𝑡_31_. We modeled 𝑅(𝑡) as a product of the secondary attack risk (i.e., risk of infection per sexual encounter) and the mean number of sexual partners of individuals infected at time 𝑡 minus one (i.e., excluding the partner who infected the case).

Here, the sexual partnership distribution from the UK National Survey of Sexual Attitudes and Lifestyles (Natsal) data ^22,32^ was assumed to apply to all included countries. The susceptible proportion among contacts is represented by 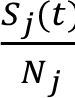, which implies random mixing apart from individual variation due to the degree distribution, i.e., no degree assortativity. The reduction in 𝑅(𝑡) over time due to infection-derived immunity has been characterized in our recent study^6,32^ (see *Supplementary Materials*).

We then modeled the importation hazard for country 𝑖 located in region 𝑔 since the start of the current global mpox outbreak (i.e., the symptom onset date of the initial case in the UK; 17 April 2022). Here, we assumed that all mpox importation events reported in country 𝑖 represent residents who returned from international travels, during which they acquired infection in a source country 𝑗. This assumption is based on our understanding that travel-related cases are highly likely to be reported in their country of origin rather than in the travel country due to diagnostic delays. The importation hazard rate in country 𝑖, *ℎ_i_*, is defined as:

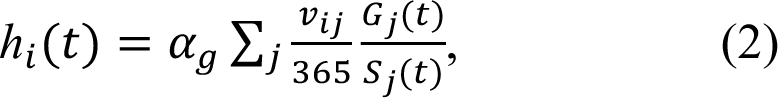

where 𝛼_+_ is a scaling factor accounting for the reporting probability and the likelihood of engaging in sexual activity while traveling abroad, which we varied between six regions (i.e., Africa, Americas, Asia, Europe, Middle East, and Oceania; defined by the UN geoscheme ^33^) to represent possible regional-level effect. The annual travel volume 𝑣_*ij*_ from country 𝑗 to 𝑖 was assumed to be constant throughout the year. 𝑆_*j*_ is the susceptible population size in country 𝑗. As we only consider the first importation event in country 𝑖, returning travelers from country 𝑗 to 𝑖 are assumed to be susceptible and thus their risk of becoming a case is given as 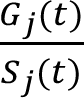. The survival probability of country *i* experiencing no importation event by time 𝑡 is given by

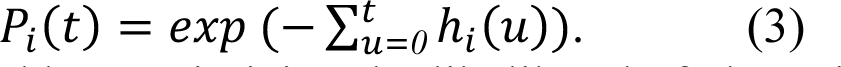

Parameters 𝛼_+_ were estimated by maximizing the likelihood of observing the first importation event in each country, with 95% confidence intervals derived from the likelihood ratio. Further details on the parameter estimation are presented in *Supplementary Materials*.

### Model selection & Counterfactual analysis

We used a model selection approach to assess the support for the following hypotheses given the observed data: (1) the importation hazard in each country is explained only by the temporal changes in the number of cases in source countries weighted by travel volume, (2) extra reduction reflecting selective depletion needs to be accounted for, and (3) including geographical heterogeneity in the importation hazard model better describes the data. To this end, we compared a total of four different models, with and without selective depletion and using the global and region-stratified scaling factors. In the models without selective depletion, 𝑅(𝑡) was assumed to be constant; the cumulative cases in affected countries only account for a negligible proportion (typically < 1%) of the estimated MSM population^6^, which is unlikely to affect 𝑅(𝑡) if the transmission dynamics is homogeneous (i.e., non-selective). In the models with the global scaling factor, a single parameter 𝛼_+_ = 𝛼 was used across all regions. The best model was selected based on the Laplace-approximated model evidence (see *Supplementary Materials*).

To illustrate the impact of the selective depletion effect, our model with selective depletion was compared with the counterfactual model that assumes no network heterogeneity (thus no selective depletion). The two models were assumed to share the same initial reproduction number, and the temporal changes in the regional average of the importation hazard in both models were displayed.

### Export capacity

Based on the selected model, we estimated the export capacity, which represents a country’s remaining potential to export mpox cases to other countries at a given time. We defined the export capacity as the number of cases that a country is capable of exporting in theory by the end of an epidemic in the absence of any interventions or behavior changes. The export capacity of country 𝑖 at time 𝑡, 𝐶_*i*_(𝑡), is expressed as

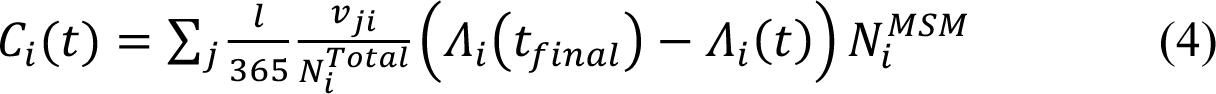

where 𝑙 is the mean duration of international trips (assumed to be 7 days). *𝑁^MSM^_i_* and *𝑁^Total^_i_* are estimated MSM and total population size in country 𝑖. 𝛬_*i*_(𝑡) and 𝛬(*t_final_*) represent the cumulative force of infection at a given time 𝑡 and the time at the end of an epidemic, which is projected by an mpox transmission model (assuming no intervention or behavioral changes) used in our previous study^6^. 𝛬_*i*_(*t_final_*) was computed based on the identical secondary attack risk across countries (assumed to be 0.2), while we conducted a sensitivity analysis with different levels of secondary attack risk (varying from 0.2 to 0.4).

## Results

Mpox importation events in the current outbreak were observed first in Europe, followed by North America, the Middle East, and other regions (**Fig-1**). The cumulative curve for importation by reporting date shows an apparent swift surge in mid-May (**Fig-1A**). However, this is likely due to reporting delays (including initially unrecognized cases) and is smoothed out in the curve by date of onset (**Fig-1B**). A steeper slope was observed between May and June, and the flatter curve after August indicated the saturating trend in the rate of the first importation event. The pattern of mpox international spread in relation to the increase in instantaneous case counts was clearly distinct from that of COVID-19 despite both diseases sharing similar initial estimates of R_0_ ∼ 2^12,13^ (**Fig-S1 & S2**). The increase in the number of countries that have ever imported COVID-19 suggests an acceleration in the rate of importation, consistent with the growth of the global case counts over time (**Fig-S1(B)** & **S2(B)(D)**). In contrast, many mpox importations were observed when the number of cases was much smaller compared with COVID-19 but the rate of international spread quickly saturated (**Fig-S1(A) & S2(A)(C)**).

**Figure 1.**
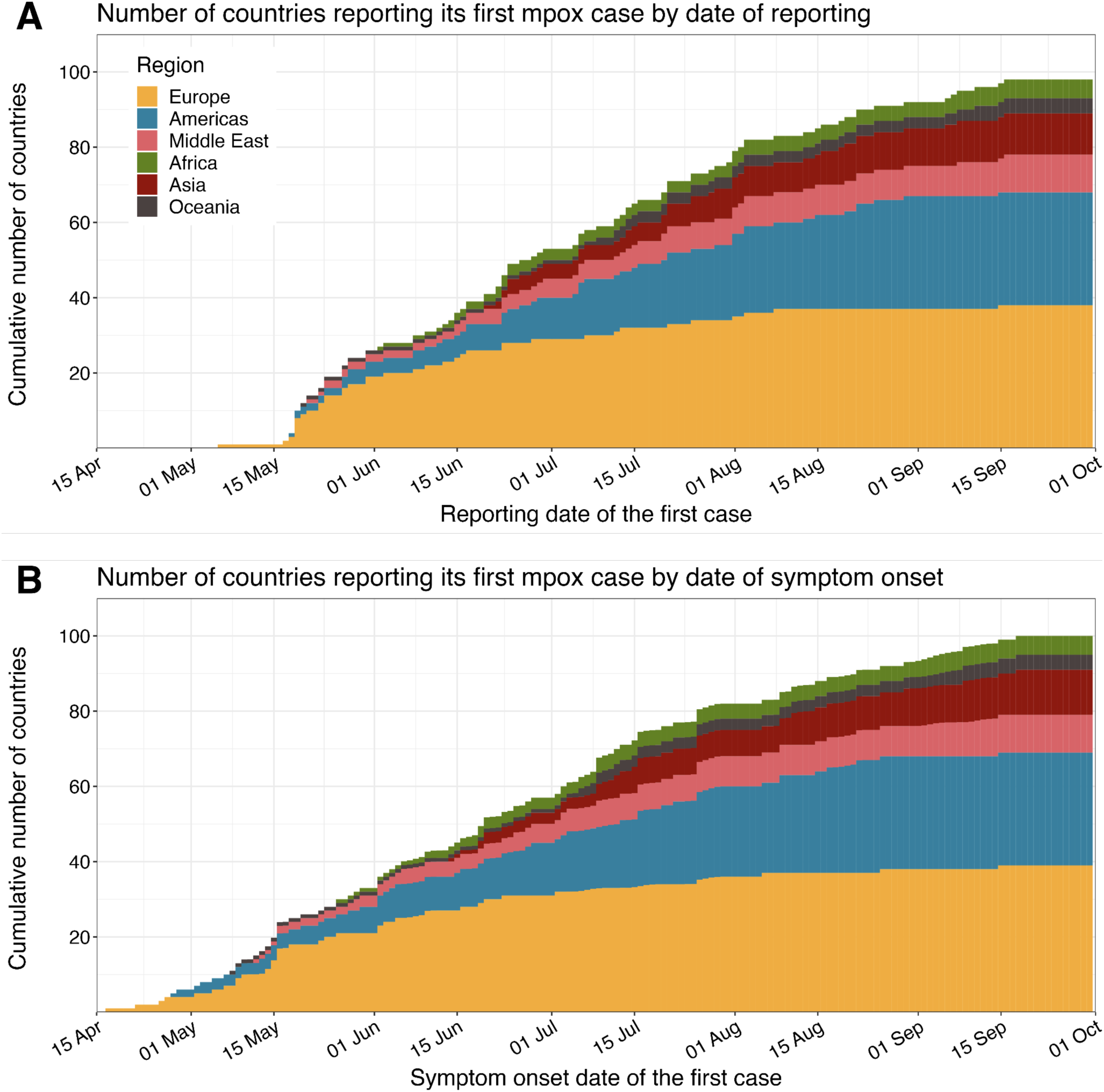
Number of countries reporting its first mpox case. The cumulative number of countries reporting its first mpox case by **(A)** date of reporting and **(B)** date of symptom onset. The regions that each country is located in are shown by colors. When the symptom onset date of the first case is unavailable, its imputed distribution was used to show all potential dates.

Our proposed model accounting for the selective depletion effect explained the observed trend of mpox importations better than the models without selective depletion (**Table-S1**). Out of the models that incorporated selective depletion, the model with region-specific scaling factors was selected as the best model, suggesting a significant variation in the importation hazard between regions. The goodness-of-fit of the selected model was visualized by comparing the modeled cumulative importation hazard by the first importation event in each country (represented by a straight line on a semi-log plot; **Fig-S3**). The observed coverage of our (in-sample) 95% prediction interval for the first importation dates was 80%, suggesting that the model fit was overall good but not perfect (**Fig-S4**).

The estimated hazard of importation by country showed the global distribution of mpox importation risks over time (**Fig-2**). Throughout the included period, many countries in Europe were estimated to have experienced particularly high hazards of importation, which explains their early importation dates. By contrast, the estimated hazard of importation was substantially low for countries with late importation events (e.g., countries without the first importation by the end of October 2022). Such geographical variation was highlighted in the time series of regional-average importation hazard (**Fig-3**). The importation hazard was highest in Europe throughout the analyzed period until October while it was 10-30 times lower in Asia, Oceania, the Middle East, and Africa. We also displayed the modeled importation hazards in the counterfactual scenario where the selective depletion effect due to sexual network heterogeneity was assumed to be negligible (**Fig-3 & S5**). The results highlighted the importance of selective depletion in explaining the decline in importation events.

**Figure 2.**
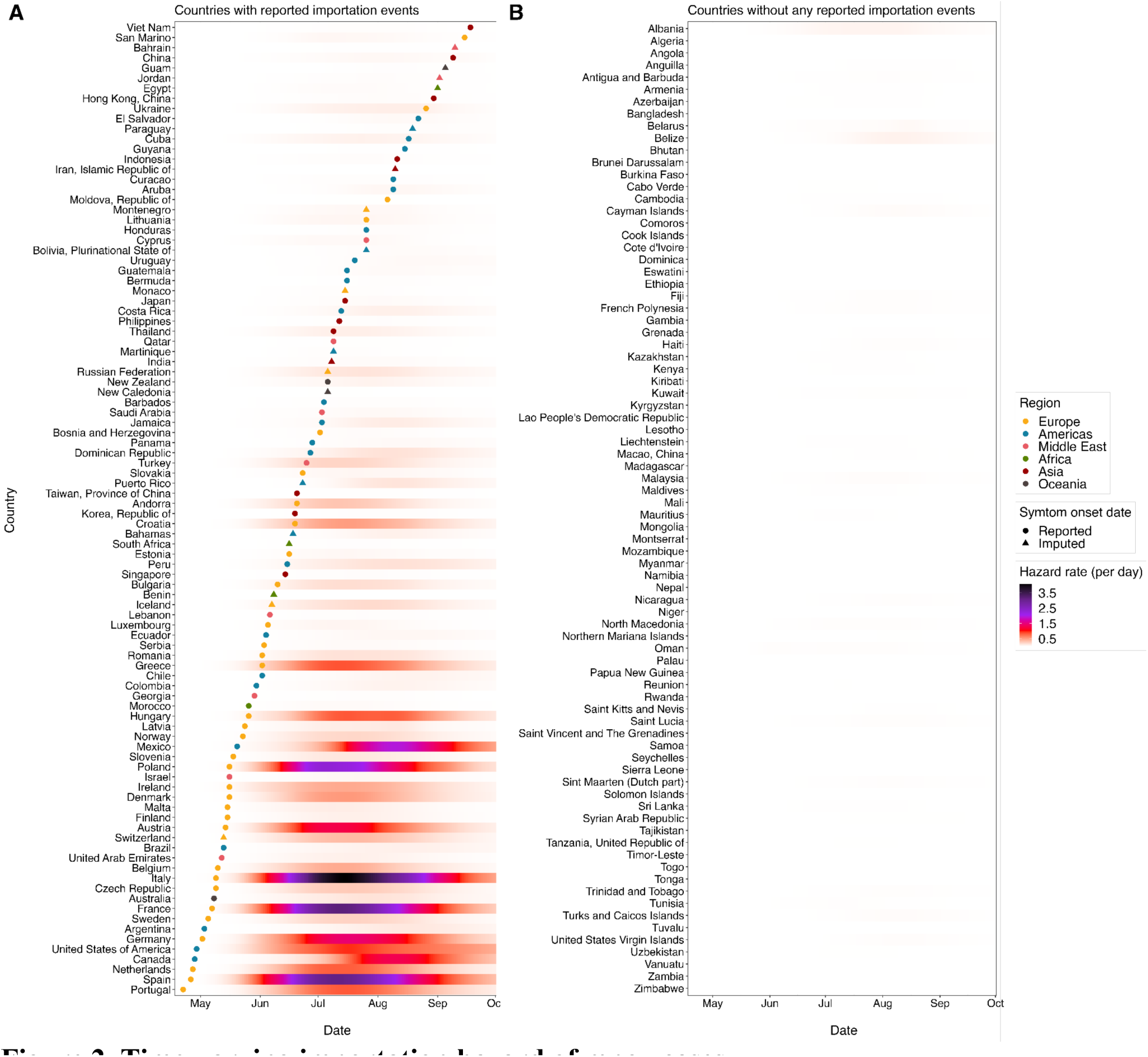
Time-varying importation hazard of mpox cases. Estimated time-varying importation hazard of mpox in **(A)** countries have reported mpox importation events and **(B)** countries that have not, as of 1 October 2022. The best model accounting for selective depletion effect and region-specific scaling factors was applied to compute the time-varying importation hazard, and it was shown as a color scale. Dots and triangles indicate the reported and the median of the imputed symptom onset dates of the first confirmed mpox case in each country, respectively. The colors of dots and triangles represent the regions in which each country is located.

We retrospectively estimated the export capacity of each country at different epidemic phases (**Fig-4**). The estimated export capacity was initially high among countries with larger MSM population sizes and higher rates of international travel. As the epidemic progressed, the export capacity declined in the most affected countries, particularly in North America and Europe (**Fig-S6**), reflecting the accumulated infection-derived immunity among individuals with a higher number of sexual partners. By contrast, some countries reporting a relatively small number of cumulative cases have retained substantial export capacities at the time of analysis. In our sensitivity analysis, such temporal changes in the export capacity following the progression of outbreaks were suggested to be clearer with a lower secondary attack risk (**Fig-S7**) since the effective cohort reproduction number would rapidly fall below the value of one^6,32^.

## Discussion

The global outbreak of mpox in 2022 was first recognized in a small number of countries in Europe, followed by rapid importation events to other countries and regions in the earlier phase but subsequently much fewer in the later phase. Such a distinctive international spreading pattern of mpox was best explained by our model that incorporated both international mobility of infected individuals and selective depletion of susceptibles driven by the heavy-tailed nature of sexual contact patterns, compared to models not accounting for heterogeneous sexual contacts. Based on the proposed model and empirical data observed in 2022, we retrospectively assessed the time-varying hazard of mpox importation events and export capacity in each country, informing the landscape of global importation risk and the potential resurgence of mpox across countries.

Transmission dynamics of mpox in 2022 outbreak was likely substantially influenced by the heavy-tailed nature of MSM sexual networks^6,32^. Our analysis adds to these previous findings and suggests that the substantial heterogeneity in transmission may have also shaped the international spread patterns of mpox that appear distinct from respiratory infection outbreaks such as COVID-19. In the presence of selective accumulation of infection-derived immunity, not only do infections among the MSM population quickly saturate but also cases have increasingly less risk of onward transmission since those infected in the later phase of an epidemic tend to have lower risk profile (i.e., have fewer sexual partners as was observed^34^). The time-varying hazard of case importation, where travelers return infected from their travel destinations, would therefore decline faster than the number of cases does in the destinations. Meanwhile, such temporal patterns may be less relevant to respiratory infection outbreaks where face-to-face contact is the primary mode of transmission; heterogeneity in respiratory infections is typically represented by non-heavy-tail distributions (e.g., the negative-binomial distribution^35^), where the relative role of the tail part is smaller. We can observe the difference potentially reflecting this in the importation patterns between mpox and COVID-19: diseases that share similar initial estimates of *R*_0_ ∼ 2^12,13^ (**Fig-S1 & S2**). Such a unique pattern of mpox spread against COVID-19 may imply the importance of integrating selective depletion into the global spread model where a highly heterogeneous sexual network plays a role.

While modeling studies have been overall successful in quantifying the international spread of respiratory infectious diseases such as influenza^18^ and COVID-19^10^, directly transporting these models to the current mpox outbreak driven by sexually-associated contacts without accounting for their highly heterogeneous nature may yield misleading results. Conventional models often assume the risk of importation to be proportional to the number of cases in the source country, which may have been plausible in past respiratory infection outbreaks that exhibit relatively moderate heterogeneity^36^. Our finding, however, suggests that this assumption of proportionality may not be the optimal choice for sexually-associated outbreaks such as mpox, where empirical importation patterns were better described by the selective depletion model. In such outbreaks, the proportionality model would overestimate the risk of importation from a country where the highest risk groups have already been selectively immunized. As a result, global resources for control may be misallocated among countries—i.e., countries with a more recent epidemic onset, which have ongoing infections among the most sexually-active individuals and thus are more likely to contribute to the onward global spread, may not be prioritized over countries that have already passed that phase and are no longer exporting many cases.

Our retrospective analysis of export capacity visualized countries capable of contributing to further international spread if they experience sustained mpox transmission. The potential risk of exporting mpox cases from one country is shaped by the proportion of high-risk individuals who remain susceptible, the MSM population size, and the connectivity with other countries. Accordingly, the countries initially affected by the current outbreak showed a gradual decline in export capacity following sustained local transmissions, whereas less affected countries with sizeable susceptible populations (e.g., Asian countries) have retained a substantial export capacity (**Fig-3**). Such maldistributed risks of exporting cases highlight the importance of immunizing high-risk individuals in less affected countries to prevent the re-emergence of the global spread of mpox. However, it needs to be noted that our estimated export capacity represents the upper bound risk of exporting cases from a source country, as the final size in each country (which determines the export capacity) was modeled assuming no interventions or behavior changes. Moreover, regardless of the extent of export capacity, the likelihood of causing major outbreaks in destination countries will be determined by their own epidemiological situation such as the proportion of remaining susceptibles as well as their risk awareness. Nevertheless, our results illustrate the geographical distribution of remaining risks for mpox exportations and offer guidance for effective allocation of international control efforts.

**Figure 3.**
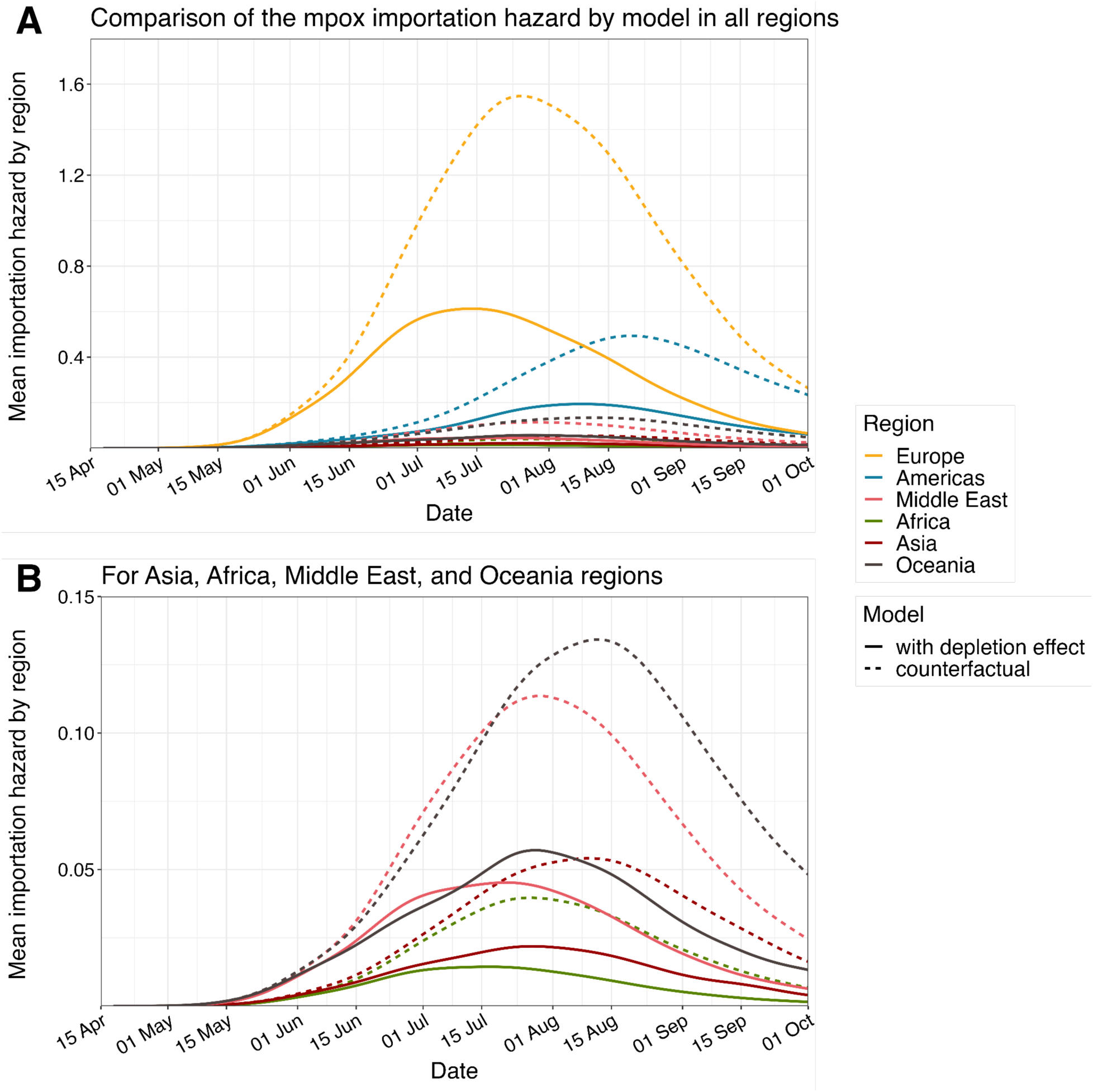
Comparison of the regional-average importation hazard of mpox cases by model. Time-varying regional-average importation hazards of mpox cases in **(A)** all regions and **(B)** less affected regions (Asia, Africa, Middle East, and Oceania). Solid lines are the fitted importation hazard using the best model accounting for selective depletion effect and region-specific scaling factors. Dashed lines are the modeled one in the counterfactual scenario where the sexual network heterogeneity was assumed to be negligible (no selective depletion).

**Figure 4.**
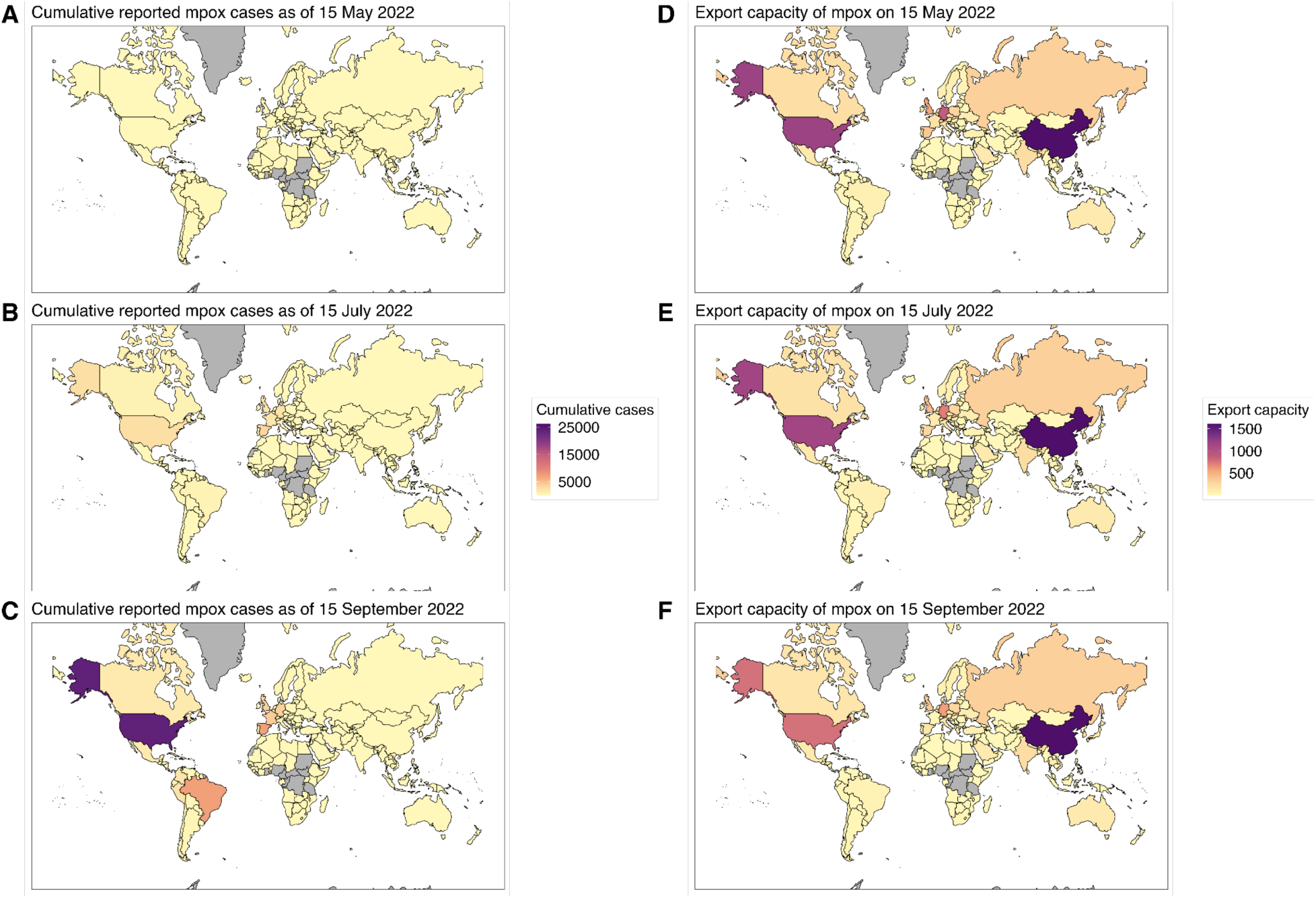
Cumulative reported mpox cases and the estimated export capacity. **(A–C)** Cumulative number of reported mpox cases and **(D–F)** the estimated export capacity with a 2-month interval from 15 May to 15 September 2022. Countries with grey coloring are either those where mpox cases existed prior to the current global outbreak or those where international travel volume data was unavailable in UNWTO 2019 outbound tourism data.

Our analysis suggests that the importation risk of mpox declined globally by the end of the study period (1 October 2022), probably even more rapidly than the case count itself because of selective depletion. However, this result does not indicate that the risk of mpox importations continues to be low in the future. Once sustained local transmissions are established in a previously less affected country where the majority of high-risk populations remain susceptible, the importation risk in other countries may (re-)surge, especially if they are closely connected in the international travel network. In fact, more than 100 local mpox cases have been reported in Japan since early 2023^37^, followed by importations and local transmissions in some of the neighboring regions (e.g., South Korea, Hong Kong, Taiwan, and China^15,38–40)^, and this may change the landscape of importation risk in other Asian countries onward. This is particularly concerning because most of these countries had been least affected by mpox in 2022 and their access to vaccines is still limited^14^, leaving a large export capacity in Asia. The set of countries at the highest ongoing risk of importation would change over time, depending on the evolving situation of the mpox outbreak. Furthermore, such resurgence could also affect the countries that experienced mpox outbreaks in early 2022, if vaccine- and infection-acquired immunity against mpox wane rapidly over time^41^.

Several limitations must be noted. First, we assumed that the sexual partnership distribution estimated for the MSM population in the UK^22^ was applicable to all included countries throughout the study period. We also assumed that mpox transmission is almost completely restricted to the MSM population, reflecting the significant role of the MSM population in the current mpox dynamics^4^. Our estimates might change if mpox establishes itself over the network of heterosexual individuals with many sexual partners (e.g., commercial sex workers). Second, the estimated MSM population sizes are subject to uncertainties due to different years of data collection and data sources by country. Third, we assumed that the case ascertainment rate in all regions remains constant in the importation hazard model. Fourth, the actual range of the secondary attack risk in the current mpox outbreak is still largely unknown, although our sensitivity analysis suggested the robustness of our qualitative conclusions (**Fig-S7**). Lastly, the mobility of infected individuals was parameterized using the UNWTO data, which may not fully reflect the actual movement of mpox cases, if there are discrepancies arising from different travel patterns between high-risk individuals in mpox transmissions and general travelers. Besides, this data does not capture sudden changes in mobility, such as festival-associated travels, which may explain the observed mpox importations in many European countries that were earlier than our model outputs (**Fig-S4**)^1,42^. Although this travel data was the best available empirical data covering both land and flight routes at the time of analysis, more accurate and standardized data on the mobility of individuals, particularly among the MSM population, would improve the robustness of our results.

## Conclusions

Our study suggests that the accumulated immunity among high-risk individuals has contributed to a slowdown in mpox importations between countries. However, there remain large susceptible populations among less affected regions, including low- and middle-income countries, without sufficient vaccine supply. To prevent future outbreaks and potential global resurgence of mpox, it is crucial to ensure equitable access to treatment and control measures in these countries at greater risk.

### Contributors

AE conceptualized the study. S-mJ, FM, and AE designed the model. S-mJ and FM contributed to data compilation and curation. S-mJ and HM contributed to data analysis, and S-mJ made figures. FM and AE consulted on the code. S-mJ, FM, and AE contributed to data interpretation and the initial manuscript draft. All authors read and approved the final version for submission.

### Data sharing

The aggregated and case-based data has been available on the WHO website^4^ and the estimated MSM population size data has been available on the UNAIDS dashboard and report^26,27^. The replication codes and all utilized data, except for the international travel volume data which was obtained from UNWTO^29^, are available at: https://github.com/SungmokJung/mpox_global.

## Data Availability

The aggregated and case-based data has been available on the WHO website and the estimated MSM population size data has been available on the UNAIDS dashboard and report. The replication codes and all utilized data, except for the international travel volume data which was obtained from UNWTO, are available at: https://github.com/SungmokJung/mpox_global.

## Acknowledgement

S-mJ and JL are supported by the Centers for Disease Control and Prevention (CDC) SHEPheRD (200-2016-91781). FM is supported by JSPS Grants-in-Aid KAKENHI (20J00793 and 21KK0107). FM and JW received funding from the European Union’s Horizon 2023 research and innovation programme - project ESCAPE (Grant agreement number 101095619). AE is supported by Japan Society for the Promotion of Science (JSPS) Overseas Research Fellowships, JSPS Grants-in-Aid KAKENHI (22K17329), foundation for the Fusion Of Science and Technology, and Japan Science and Technology Agency (JST) Precursory Research for Embryonic Science and Technology (JPMJPR22R3). This research was supported by Japan Agency for Medical Research and Development (AMED) under Strategic Center of Biomedical Advanced Vaccine Research and Development for Preparedness and Response (SCARDA). The funders had no role in study design, data collection and analysis, decision to publish, or preparation of the manuscript.

## *Supplementary Materials*: Dynamic landscape of mpox importation risks driven by heavy-tailed sexual contact networks among men who have sex with men: a mathematical modeling study

### Global mpox incidence

When symptom onset dates of the reported mpox cases were unavailable, they were reconstructed from the reporting date using the estimated reporting delay distribution. In detail, we estimated time delays from the symptom onset to report using empirical data from countries that met the WHO reporting quality criterion, gathered from the WHO website as of 1 October 20204. Given cases where no time delays were reported (accounted for 18% of the included cases), a zero-inflated discrete lognormal distribution was fitted to the data (mean of 8 days with SD of 6 days). We then performed a non-parametric back-projection using the parameterized discrete delay distribution and R package ‘*surveillance*’^43^. We also smoothed the incidence curve by taking a 14-day moving average to minimize the daily noise. Case data reported to the WHO as China was stratified into three (Hong Kong, Taiwan, and mainland China) using the official reports^39,40^.

### International travel volume

International travel volume was available in the UNWTO 2019 outbound tourism data^29^. The outbound travel volume was defined as “the annual number of international trips by resident visitors of each country to destination countries”, which included eight data series based on the arrivals of non-resident visitors with slightly different definitions of ‘visitors’ (by nationality or country of residence), the main purpose of travel (tourism only or all-purpose travels), and collection sites (at national borders, hotels, or all types of accommodations). Since the availability of these series varied by country, we selected the maximum value among the available series for each country.

### Global COVID-19 incidence

We obtained information on global COVID-19 cases from the WHO website^44^. The reporting date of the first case in each country (arrival time) was validated against external sources (e.g., official reports from governmental institutes) and was adjusted if the WHO data were outdated. This modification was carried out until ten countries consecutively showed a difference of less than two days (given the time difference between countries). The arrival time of COVID-19 along with the utilized sources is provided as *Supplementary Material 2*.

### Reproduction number

We modeled the time-varying reproduction number (𝑅(𝑡)) of mpox by accounting for the selective depletion of susceptibles over highly heterogeneous sexual contact networks. We assumed the distribution of the number of sexual partners over the infectious period of mpox (assumed to be 14 days) in included countries is represented by a Weibull distribution, previously parameterized based on Natsal data^22,32^. We then modeled 𝑅(𝑡) following our recent study^6^, as a product of the secondary attack risk and the mean excess degree^45^ among currently infected individuals (i.e., the number of exposed partners per infected individual). Here, we assumed that all individuals in a country were fully susceptible to mpox initially and the risk of contracting mpox was proportional to the number of sexual partners. We also assumed that infected individuals develop long-term immunity upon recovery and keep their sexual behavior without the risk of reinfection.

### Likelihood function for estimating parameters

The likelihood function with regard to the date of first importation in the included countries is formulated as a survival process:

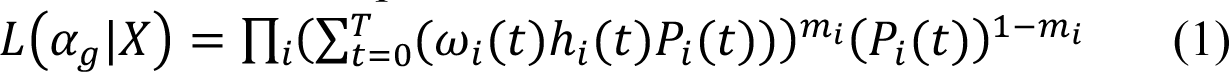

where 𝑋 is the observed data containing the daily cumulative cases for all countries from 𝑡 = *0* (i.e., the symptom onset date of the initial case in the UK) to 𝑇 (i.e., the end of the included period). Here, the UK was excluded from the observed data 𝑋 since the initial case reported in the UK was assumed to be the first case of the current global mpox outbreak in our study. 𝑚_*i*_ is a dummy variable for right censoring (𝑚_*i*_ = *1* for countries with importation events, 𝑚_*i*_ = *0* for countries that have not yet reported any mpox cases throughout the included period). We introduced 𝜔_*i*_(⋅), the probability mass function of the symptom onset date of the first imported case in country 𝑖 (derived from empirical data and the reconstructed dates via back-projection), to marginalize out the unknown onset dates when the exact date of symptom onset is not available. When the symptom onset date is known, 𝜔_*i*_(⋅) is a Kronecker delta: 𝜔_*i*_(𝑡) = 𝛿_$’7”_, where *T_i_* is the known date of onset for the first imported case in country *i*. The maximum likelihood method was employed and 95% confidence intervals were computed using the likelihood ratio.

### Laplace-approximated model evidence

In our study, a total of four models incorporating different combinations of (1) the depletion effect (with and without) and (2) scaling factors (global and region-specific) were considered. To determine which model best describes the observed mpox importation patterns, we compared the four candidate models and selected the best model by the Laplace-approximated model evidence (LAME)^46,47^, which is closely related to the Bayesian Information Criterion (BIC) but does not rely on the assumption that data are independent and identically distributed (i.i.d). Given the non-independent nature of our data where the occurrence of importation events in a country would affect the importation hazard in other countries, we employed the LAME which is defined as:

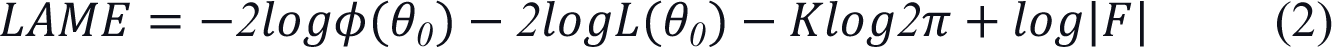

where 𝜙(𝜃*_0_*) is the prior density of the maximum a posteriori estimate, for which we used an improper flat prior (𝜙(𝜃) = *1*). 𝐿(𝜃*_0_*) is the total likelihood at the maximum a posteriori estimate. 𝐾 is the parameter dimension and |𝐹| is the determinant of the Fisher information matrix. When the LAME of a model is smaller than any other model by at least 4 ^48^, the model was selected as the best model (**Table-S1**). All analysis was performed in R version 4.2.2, and replication codes are available online (https://github.com/SungmokJung/mpox_global).

**Table S1.** Summary of the estimated parameters and model comparison based on Laplace-approximated model evidence. Model 1: with the selective depletion effect and global scaling factor; Model 2: with the selective depletion effect and region-specific scaling factors; Model 3: without the selective depletion effect and global scaling factor; Model 4: without the selective depletion effect and region-specific scaling factors Each value describes the estimated parameters (scaling factors 𝛼_+_) of four different models and their 95% confidence intervals derived from the likelihood ratio.

| Model | Parameter | Estimate<br>(95% confidence intervals) | Laplace-approximated<br>model evidence |
| --- | --- | --- | --- |
| Models with the depletion effect |  |  |  |
| Model 1 | $\alpha_{Global}$ | 0.0005 (0.0004–0.0007) | 1120.56 |
| Model 2 | $\alpha_{Europe}$ | 0.0013 (0.0009–0.0017) | 1044.03 |
| | $\alpha_{Africa}$ | 0.0002 (0.0001–0.0004) | |
| | $\alpha_{Americas}$ | 0.0009 (0.0006–0.0013) | |
| | $\alpha_{Asia}$ | 0.0002 (0.0001–0.0003) | |
| | $\alpha_{Middle\ East}$ | 0.0005 (0.0002–0.0008) | |
| | $\alpha_{Oceania}$ | 0.0018 (0.0006–0.0042) | |
| Models without the depletion effect |  |  |  |
| Model 3 | $\alpha_{Global}$ | 0.0061 (0.0050–0.0074) | 1152.17 |
| Model 4 | $\alpha_{Europe}$ | 0.0183 (0.0131–0.0247) | 1062.14 |
| | $\alpha_{Africa}$ | 0.0014 (0.0004–0.0033) | |
| | $\alpha_{Americas}$ | 0.0101 (0.0068–0.0143) | |
| | $\alpha_{Asia}$ | 0.0020 (0.0011–0.0033) | |
| | $\alpha_{Middle\ East}$ | 0.0059 (0.0029–0.0103) | |
| | $\alpha_{Oceania}$ | 0.0200 (0.0061–0.0455) | |

Each value describes the estimated parameters (scaling factors $\alpha_g$ ) of four different models and their 95% confidence intervals derived from the likelihood ratio.

**Figure S1.**
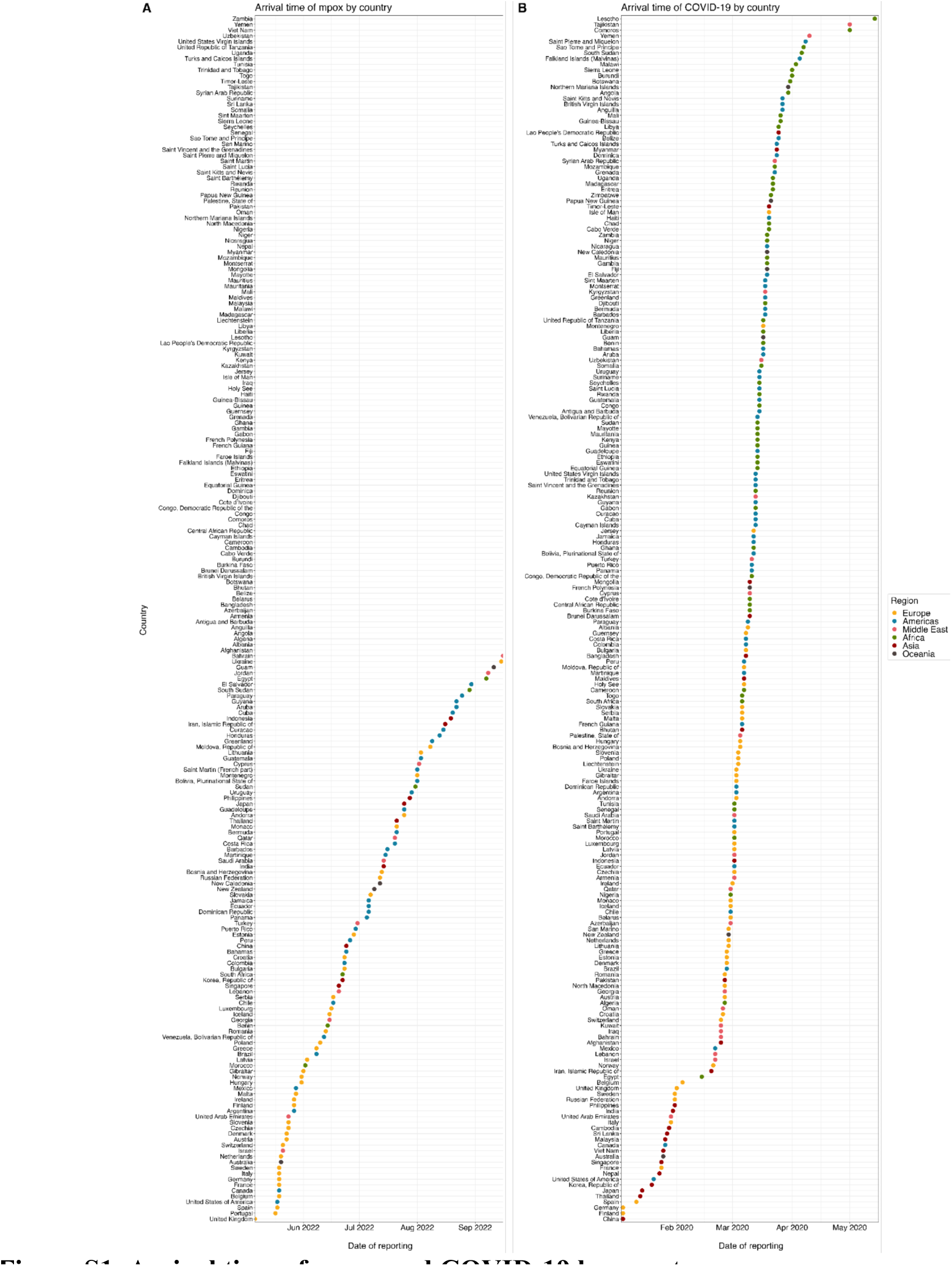
Arrival time of mpox and COVID-19 by country. Reporting dates of the first confirmed cases of **(A)** mpox and **(B)** COVID-19 in each country are shown with dots. The colors of dots represent the regions in which each country is located. **(A)** Countries without any dots are those where mpox importation events had not been reported between 6 May (i.e., the first confirmed mpox case in the UK) and 1 October 2022, while **(B)** all of the listed countries had experienced COVID-19 importation events. The same length of time (x-axis) was applied in both panels.

**Figure S2.**
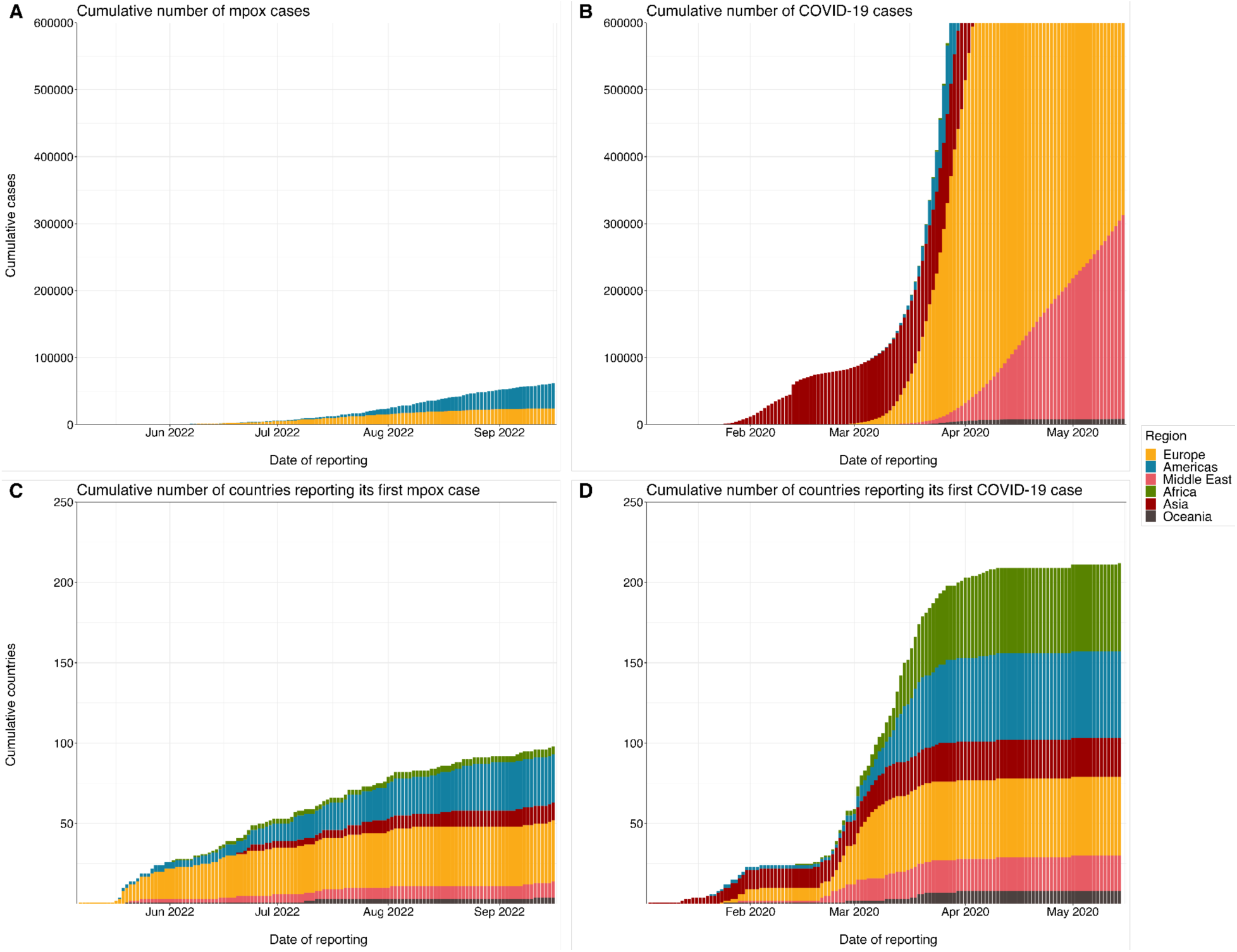
CThe cumulative number of reported cases and countries reporting its first case of mpox and COVID-19. The cumulative number of reported **(A)** mpox cases and **(B)** COVID-19 cases by date of reporting. The cumulative number of countries reporting its first **(C)** mpox case and **(D)** COVID-19 case by date of reporting. The same length of time (x-axis) was applied in all panels. The regions that each country is located in are shown by colors.

**Figure S3.**
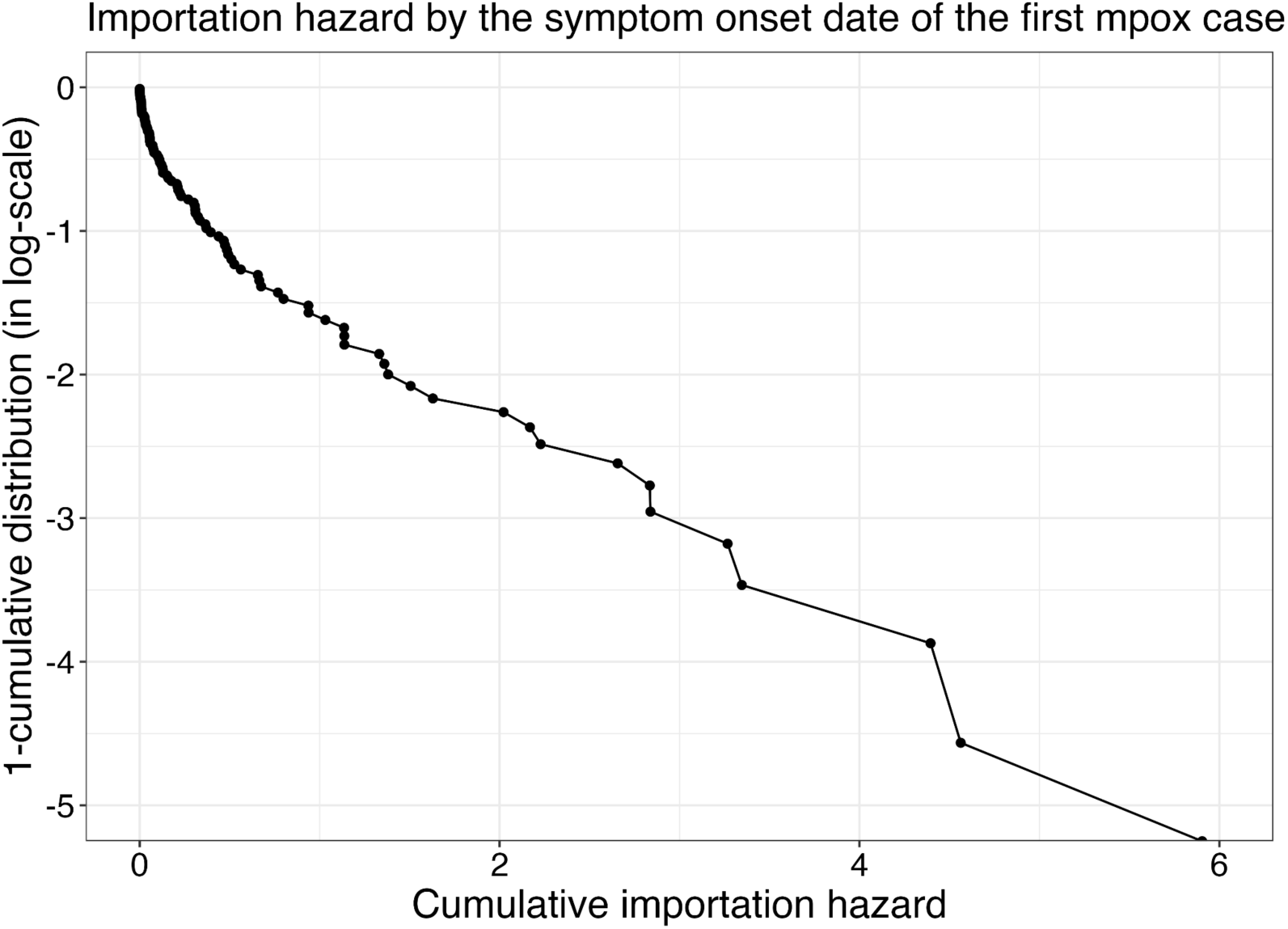
Modeled cumulative importation hazard by the first mpox importation event of each country. The cumulative importation hazard by the symptom onset date of the first importation event is plotted on the x-axis, and the survival probability for the first mpox importation event (i.e., 1 minus empirical cumulative distribution of countries with first mpox importation events) is plotted on the y-axis in the log-scale. The importation hazards were computed using the best model accounting for selective depletion effect and region-specific scaling factors.

**Figure S4.**
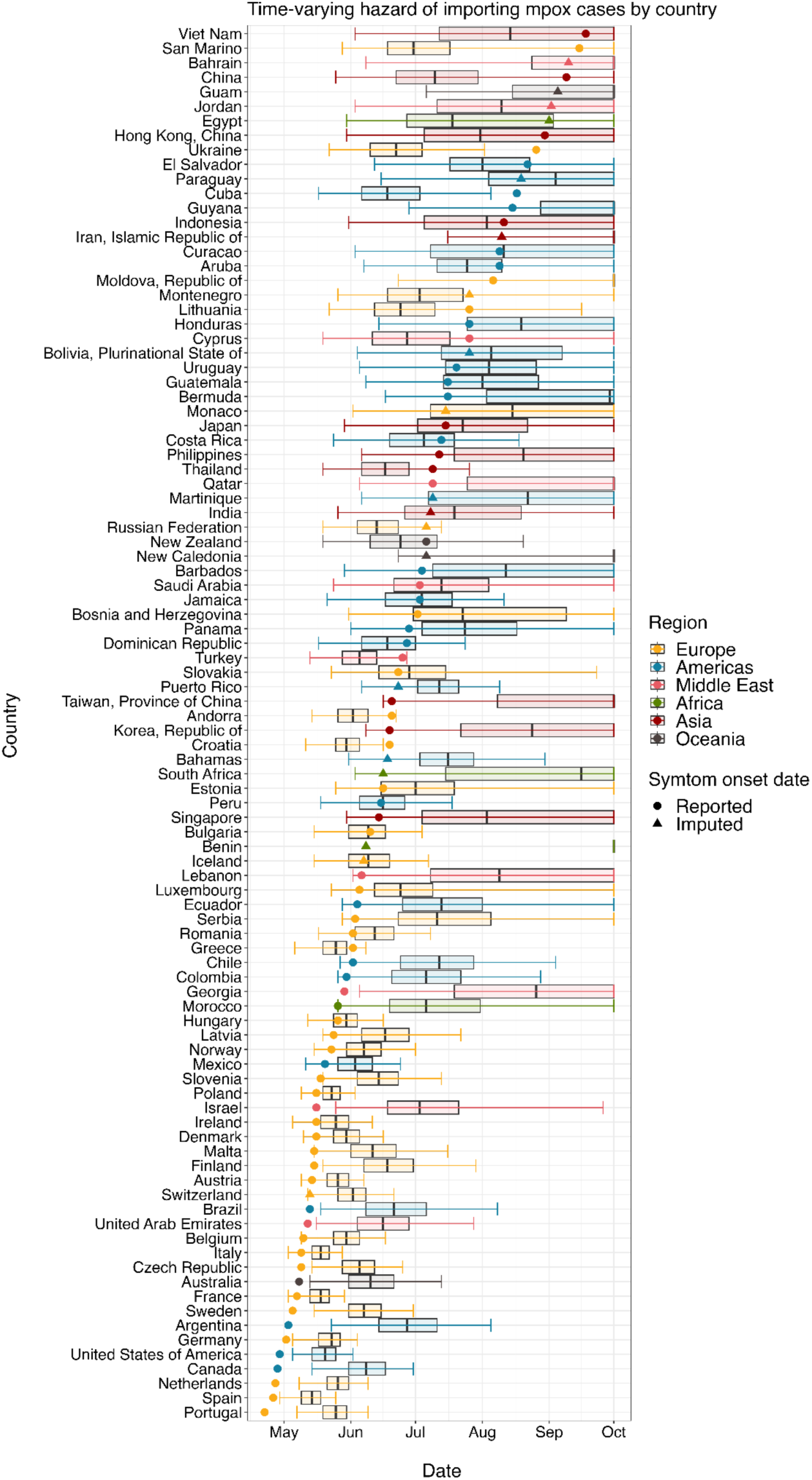
Comparison of the observed and predicted symptom onset dates of the first mpox importation event by country. Boxplots present the median and interquartile ranges of the predicted first importation date in each country, and whiskers show its 95% prediction interval. Dots and triangles are the reported and the median of the imputed symptom onset dates of the first confirmed mpox case in each country, respectively. Colors represent the regions in which each country is located.

**Figure S5.**
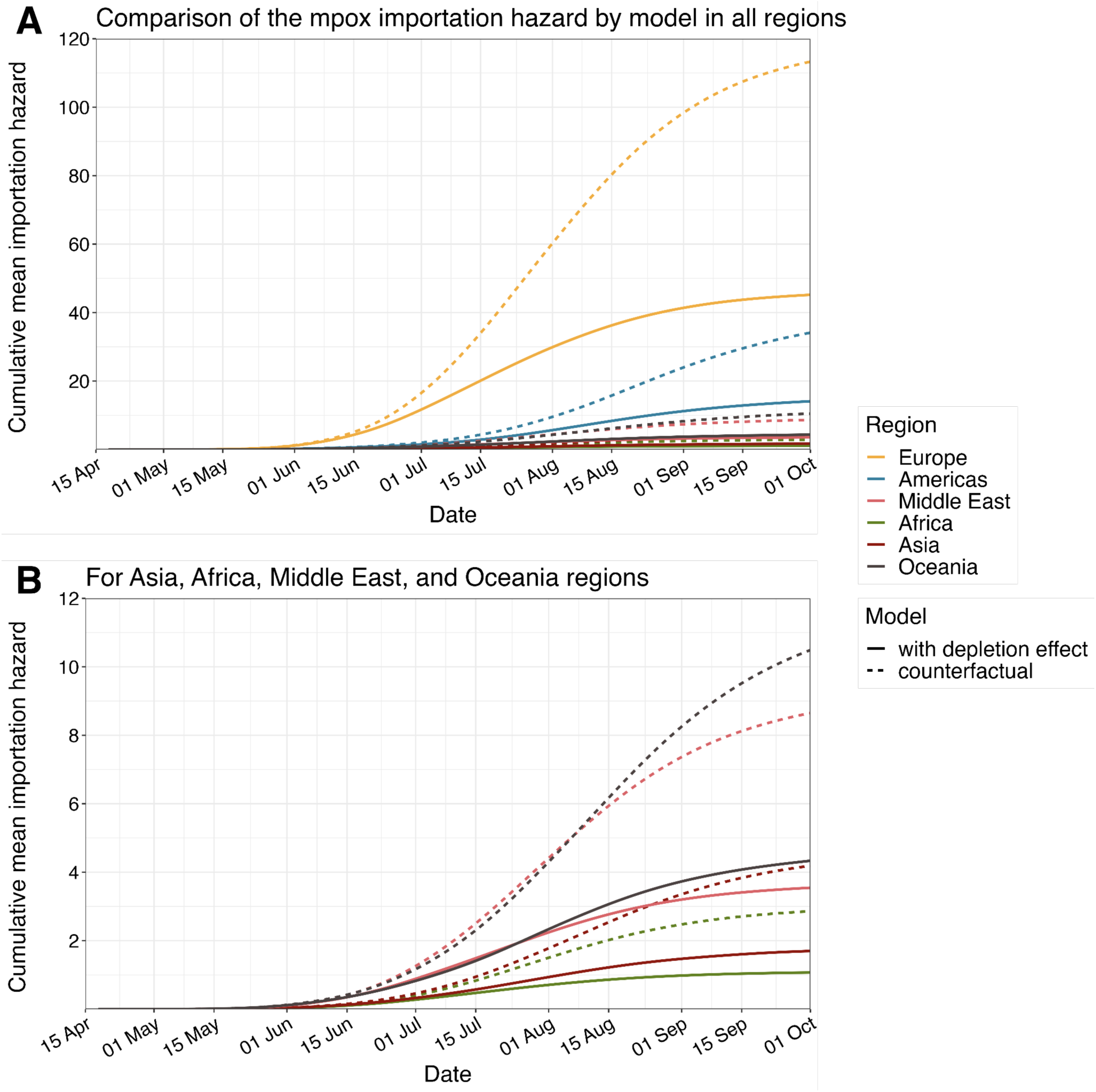
Comparison of the cumulative regional-average importation hazard of mpox cases by model. Time-varying cumulative regional-average importation hazards of mpox cases in **(A)** all regions and **(B)** less affected regions (Asia, Africa, Middle East, and Oceania). Solid lines are the fitted importation hazard using the best model accounting for selective depletion effect and region-specific scaling factors. Dashed lines are the modeled one in the counterfactual scenario where the sexual network heterogeneity was assumed to be negligible (no selective depletion).

**Figure S6.**
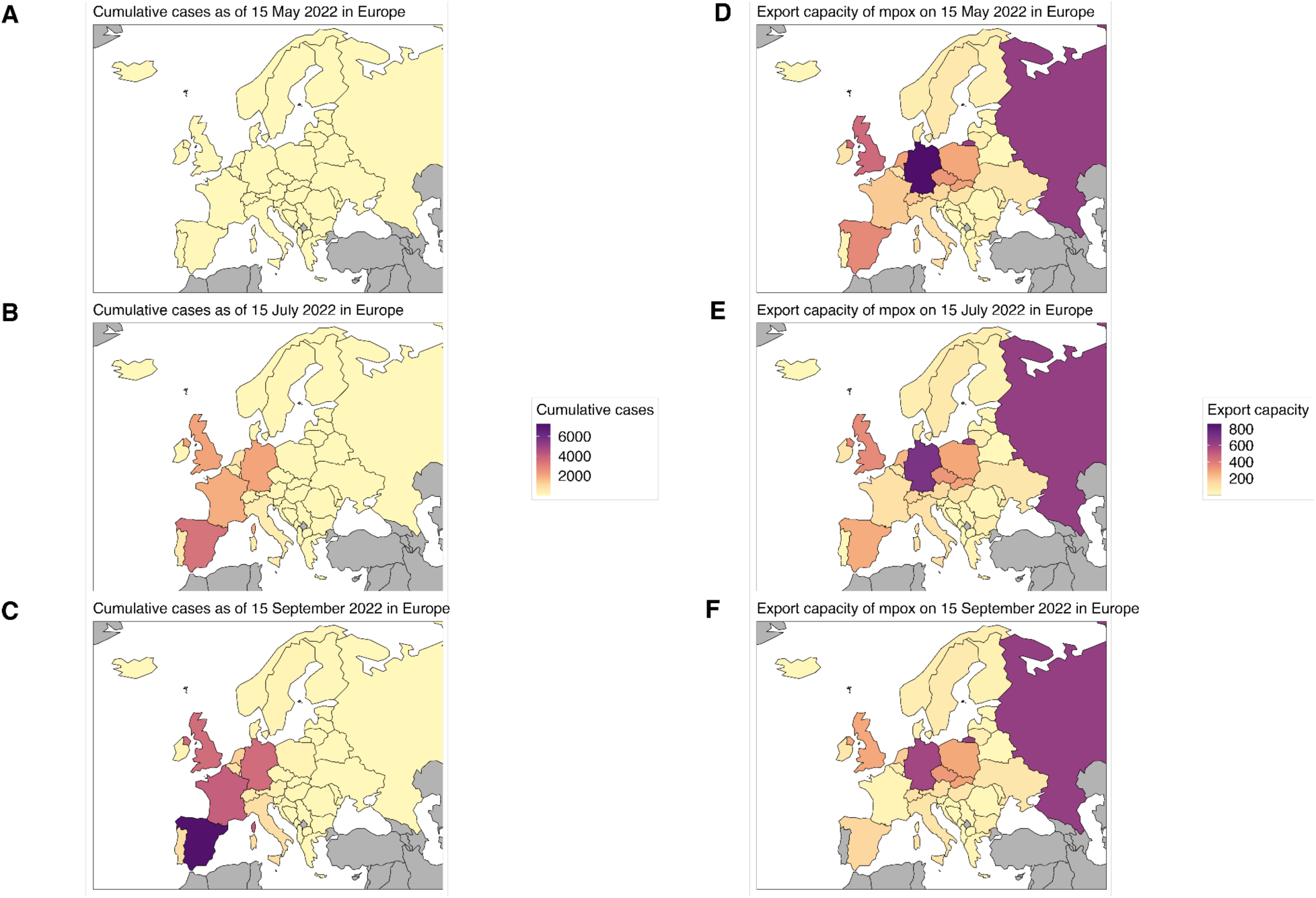
Cumulative number of reported mpox cases and the estimated export capacity in Europe. **(A–C)** Cumulative number of reported mpox cases and **(D–F)** the estimated export capacity in Europe with a 2-month interval from 15 May to 15 September 2022. Countries with grey coloring are either those not grouped in Europe region or those where international travel volume data was unavailable in UNWTO 2019 outbound tourism data.

**Figure S7.**
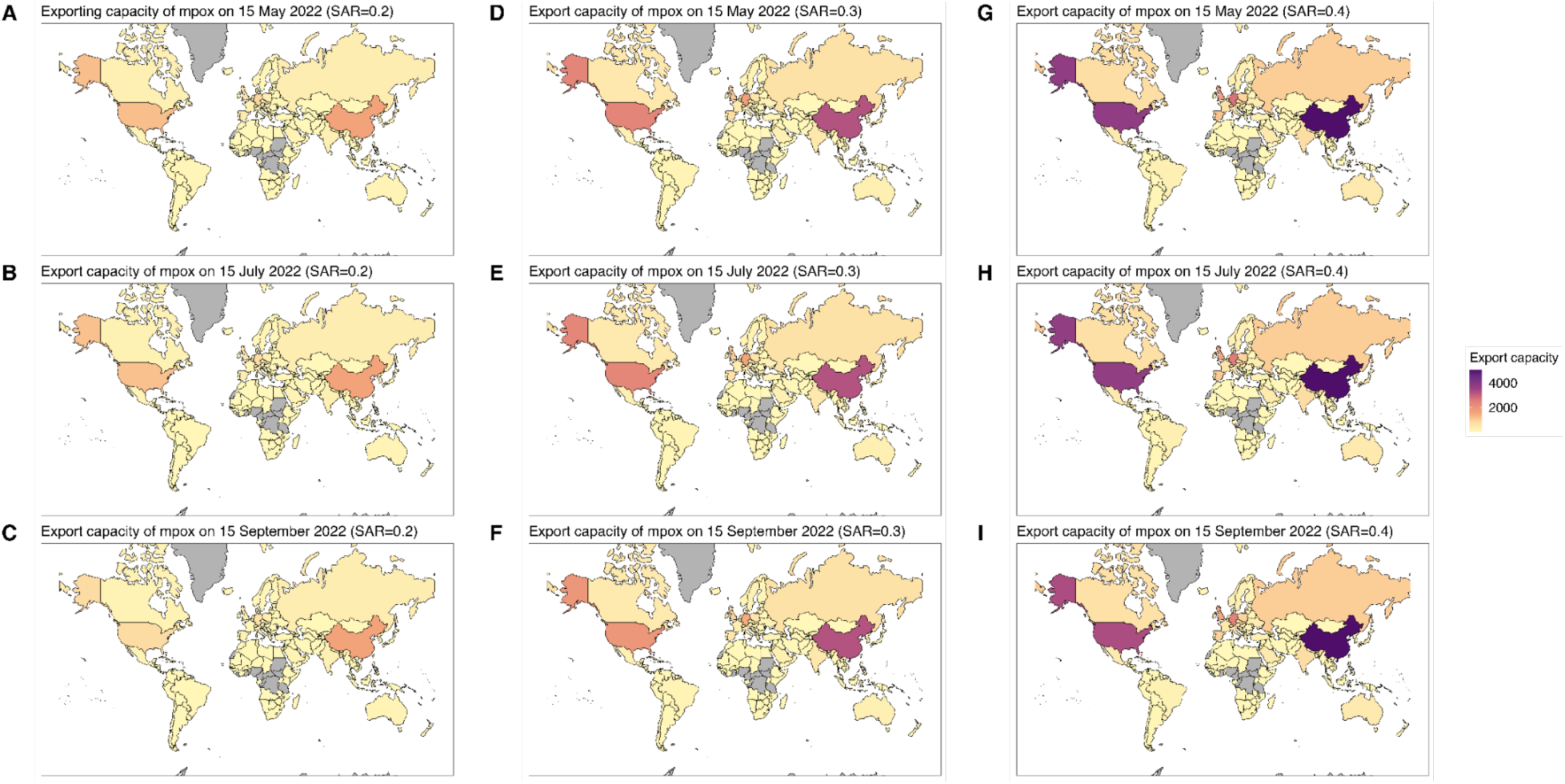
The estimated export capacity by varying the secondary attack risk of mpox. The estimated time-varying export capacity with different levels of secondary attack risk (ranging from 0.2 to 0.4) from 15 May to 15 September 2022. Countries with grey coloring are either those where mpox existed prior to the current global outbreak or those where international travel volume data was unavailable in UNWTO 2019 outbound tourism data.

